# Effects of Adapted Mindfulness Training on Interoception and Adherence to the Dietary Approaches to Stop Hypertension (DASH) Diet: The MB-BP Randomized Clinical Trial

**DOI:** 10.1101/2023.05.10.23289818

**Authors:** Eric B. Loucks, Ian M. Kronish, Frances B. Saadeh, Matthew M. Scarpaci, Jeffrey A. Proulx, Roee Gutman, Willoughby B. Britton, Zev Schuman-Olivier

## Abstract

**Background:** Hypertension is a major cause of cardiovascular disease. The Dietary Approaches to Stop Hypertension (DASH) diet lowers blood pressure (BP). However, adherence is typically low. Mindfulness training adapted to improving health behaviors that lower BP could improve DASH adherence, in part through improved interoceptive awareness relevant to dietary consumption. The primary objective of the MB-BP trial was to evaluate effects of the Mindfulness-Based Blood Pressure Reduction (MB-BP) program on interoceptive awareness. Secondary objectives assessed whether MB-BP impacts DASH adherence, and explored whether interoceptive awareness mediates DASH dietary changes.

**Methods:** Parallel-group phase 2 randomized clinical trial conducted from June 2017-November 2020 with 6 months follow-up. Data analyst was blinded to group allocation. Participants had elevated unattended office BP (≥120/80 mmHg). We randomized 201 participants to MB-BP (n=101) or enhanced usual care control (n=100). Loss-to-follow-up was 11.9%. Outcomes were the Multidimensional Assessment of Interoceptive Awareness (MAIA; range 0-5) score, and the DASH adherence score (range 0-11) assessed via a 163-item Food Frequency Questionnaire.

**Results:** Participants were 58.7% female, 81.1% non-Hispanic white, with mean age 59.5 years. Regression analyses demonstrated that MB-BP increased the MAIA score by 0.54 (95% CI: 0.35,0.74; p<.0001) at 6 months follow-up vs. control. MB-BP increased the DASH score by 0.62 (95% CI: 0.13,1.11; p=0.01) at 6 months vs. control, in participants with poor DASH adherence at baseline.

**Conclusions:** A mindfulness training program adapted to improving health behaviors that lower BP improved interoceptive awareness and DASH adherence. MB-BP could support DASH dietary adherence in adults with elevated BP.

**Clinical Trial Registration:** Clinicaltrials.gov identifier NCT03859076 (https://clinicaltrials.gov/ct2/show/NCT03859076; MAIA) and NCT03256890 (https://clinicaltrials.gov/ct2/show/NCT03256890; DASH diet adherence).

## Introduction

Hypertension is a primary cause of cardiovascular disease (CVD), and CVD is the major cause of mortality in the US and world-wide.^1^ In the United States, 47% of adults have hypertension and of them, less than 25% are controlled when defined by the 2017 ACC/AHA guidelines recommending systolic blood pressure (SBP)/diastolic blood pressure (DBP) <130/80 mm Hg.^2^ Evidence from randomized clinical trials (RCTs) and prospective observational studies shows that dietary patterns that emphasize vegetables, fruit, whole grains, lean meats, nuts and legumes, while limiting consumption of saturated fat, red and processed meats, sweets, added sugars, salt and sugar-sweetened beverages, can reduce blood pressure (BP) and incident CVD.^3,4^ While several dietary patterns are known to improve BP, evidence suggests the Dietary Approaches to Stop Hypertension (DASH) diet is particularly effective, outperforming the Mediterranean diet and caloric restriction in reducing BP.^3^

Mindfulness interventions have been tested in many diet studies, but often focused on eating disorders such as binge eating or bulimia,^5^ or weight loss.^6^ Few studies investigated the effects of mindfulness training on dietary patterns known to influence hypertension, such as the DASH diet.^6,7^ Of the mindfulness intervention studies that focused on dietary patterns, study quality is limited, follow-up time is short, and findings are mixed.^5-7^

In developing behavioral interventions, the National Institutes of Health Science of Behavior Change initiative encourages a mechanism-focus that: (1) identifies plausible mechanisms of action, (2) evaluates the impacts of a behavioral intervention on those mechanisms, and then (3) determines whether changes in the mechanisms result in behavior change.^8^ The Mindfulness-Based Blood Pressure Reduction (MB-BP) program was developed using the theoretical framework developed by Tang, Hölzel and colleagues, and adapted to CVD by Loucks and colleagues,^9,10^ focused on the mechanisms of self-awareness, attention control and emotion regulation applied to determinants of BP. It specifically trained participants in mindfulness practices using approaches grounded in the well-established Mindfulness-Based Stress Reduction (MBSR) program, and applied mindfulness training to participants’ relationships with modifiable determinants of BP including the DASH diet. No RCT has yet evaluated the impacts of MB-BP on diet or self-awareness, or evaluated self-awareness as a mediator between MB-BP and diet.

Self-awareness can be defined as awareness of one’s thoughts, emotions and physical sensations. A critical aspect of self-awareness is interoceptive awareness, which is defined as the conscious level of interoception with its multiple dimensions that are accessible to self-report.^11^ In the past decade, interoception has become more broadly understood and defined as the process of sensing, interpreting, and integrating signals originating from inside the body, including the processes of interoceptive regulation.^12^ Within the context of dietary behavior and BP regulation, interoceptive awareness may be understood as the conscious experience arising from the attention, appraisal, integration and regulation of internal sensations related to the physiological condition of the body, such as hunger and satiety cues,^13^ and noticing how different food types make patients feel.^6^

We hypothesized that interoceptive awareness is a mechanism by which MB-BP influences DASH dietary pattern, and pre-registered interoceptive awareness as the primary outcome on clinicaltrials.gov (NCT03859076). The primary objective was to evaluate effects of MB-BP on interoceptive awareness. Secondary objectives assessed whether MB-BP impacts DASH adherence, and explored whether interoceptive awareness mediates DASH dietary changes.

## Methods

### Study Design, Setting and Participants

The MB-BP Study was a parallel-group phase 2 RCT comparing group-based mindfulness meditation training adapted to improving health behaviors that lower BP versus an enhanced usual care control. Participants were recruited and enrolled from June 2017 to November 2020 through advertising in the local community and referrals from clinicians. The trial was registered with ClinicalTrials.gov at the onset of participant recruitment (applied in July 2017, approved in August 2017) and prior to outcome assessment in participants. Data were not examined prior to registration.

Inclusion criteria were: (1) elevated unattended office BP (i.e., SBP≥120 mmHg systolic or DBP ≥80 mmHg);^14^ and (2) ≥18 years of age. Exclusion criteria were: (1) Current regular meditation practice (>once/week); (2) serious medical illness precluding regular class attendance; (3) current substance use disorder, suicidal ideation or eating disorder; (4) history of bipolar or psychotic disorders or self-injurious behaviors; and (5) inability to communicate in English. The study protocol was approved by the institutional review board at Brown University (protocol #1412001171) on June 12, 2017. All participants provided written informed consent.

This study combined two unanalyzed, preregistered, clinical trials with identical methods that were part of a larger study, described elsewhere.^15^ Specifically, the study received funding through an NIH UH2/UH3 cooperative agreement that provided an initial three years of funding (UH2 phase), followed by an application for two additional two years of funding (UH3 phase). During the UH2 phase, we conducted a single arm pilot MB-BP trial, reported elsewhere.^9^ We were encouraged to shift from a single arm trial to a randomized controlled trial before the UH3 phase, which we did, preregistering the trial on clincialtrials.gov. That trial identified DASH diet and SBP at six months follow-up as the primary outcomes (#NCT03256890). The SBP findings are reported elsewhere.^15^ As we applied to the UH3 phase, we were required to create a new clinicaltrials.gov registration focused on mechanisms of behavior change. We preregistered interoceptive awareness as the primary outcome (#NCT03859076), in order to be consistent with the Science of Behavior Change model described above.^8^ We planned to combine both datasets for meta-analyses, as the clinical trial methods were identical, however we also report sensitivity analyses on interoceptive awareness for the sample preregistered in NCT03859076 that named it as the primary outcome. Limitations of this approach are described in the Discussion section.

### Blinding and Randomization

Eligible participants who completed their baseline assessments were randomized to receive either MB-BP or the control condition. Randomization was stratified within gender and BP status (i.e., SBP≥140 mmHg or DBP≥90 mmHg vs SBP 120-139 mmHg and DBP<90 mmHg). Randomization occurred approximately one week prior to the beginning of each cycle of group MB-BP classes, and participants were informed within 24 hours after randomization. Randomization was performed by a researcher blinded to participant identification using Research Randomizer (Version 4.0). The senior project manager notified participants of randomization results. Staff conducting follow-up activities were blinded to group allocation, with the exception of adverse events relatedness inquiry which was completed by a separate staff member. The data analyst was blinded to group allocation.

### Intervention Descriptions and Theoretical Framework

MB-BP is based on, and time-matched to, the standardized Mindfulness-Based Stress Reduction (MBSR) program.^9^ Both consist of a group orientation session, eight 2.5-hour weekly group sessions, and a 7.5-hour one-day group session (total of 10 sessions). Recommended home mindfulness practice was ≥45 minutes/day, 6 days/week. Adaptations to MBSR that were specific to MB-BP, described in detail in **Supplement 1** and elsewhere,^9^ were education about hypertension risk factors, along with specific mindfulness modules focused on mindful awareness of diet, physical activity, medication adherence, alcohol consumption, stress, and social support for behavior change. MB-BP builds a foundation of mindfulness skills (e.g., meditation, yoga, self-awareness, attention control, emotion regulation) through the MBSR curriculum. MB-BP directs those skills towards participants’ adhering to behaviors that can lower BP (**Figure 1; Supplement 1**).^9^ MB-BP participants have their BP and hypertension risk factors directly assessed at baseline, and are provided with this information along with the recommended best practices and national health standards for BP control. MB-BP encourages participants to explore personal readiness for change in the different hypertension lifestyle behaviors, and trains participants to utilize mindfulness practices to engage with those behaviors that they choose. Details on the diet-specific components of MB-BP are in **Supplement 1**. MB-BP was led by qualified MBSR instructors with prior expertise in cardiovascular disease etiology, treatment, and prevention. They were further trained and certified to teach MB-BP. Classes were provided in-person for the first ten cohorts (n=173), while the final two cohorts (n=28) began in-person and shifted to online via Zoom because of the COVID pandemic. Classes were held in Providence RI, USA at Brown University and at a local community health center located in a low income, urban neighborhood. Intervention group patients also received a home BP device (Omron, Model PB786N), training in home BP monitoring,^16^ and if office BP was >=140/90 mmHg, notification of primary care physicians about their BP results; those without a primary care physician were offered assistance finding a primary care physician within the constraints of their health insurance.

**Figure 1.**
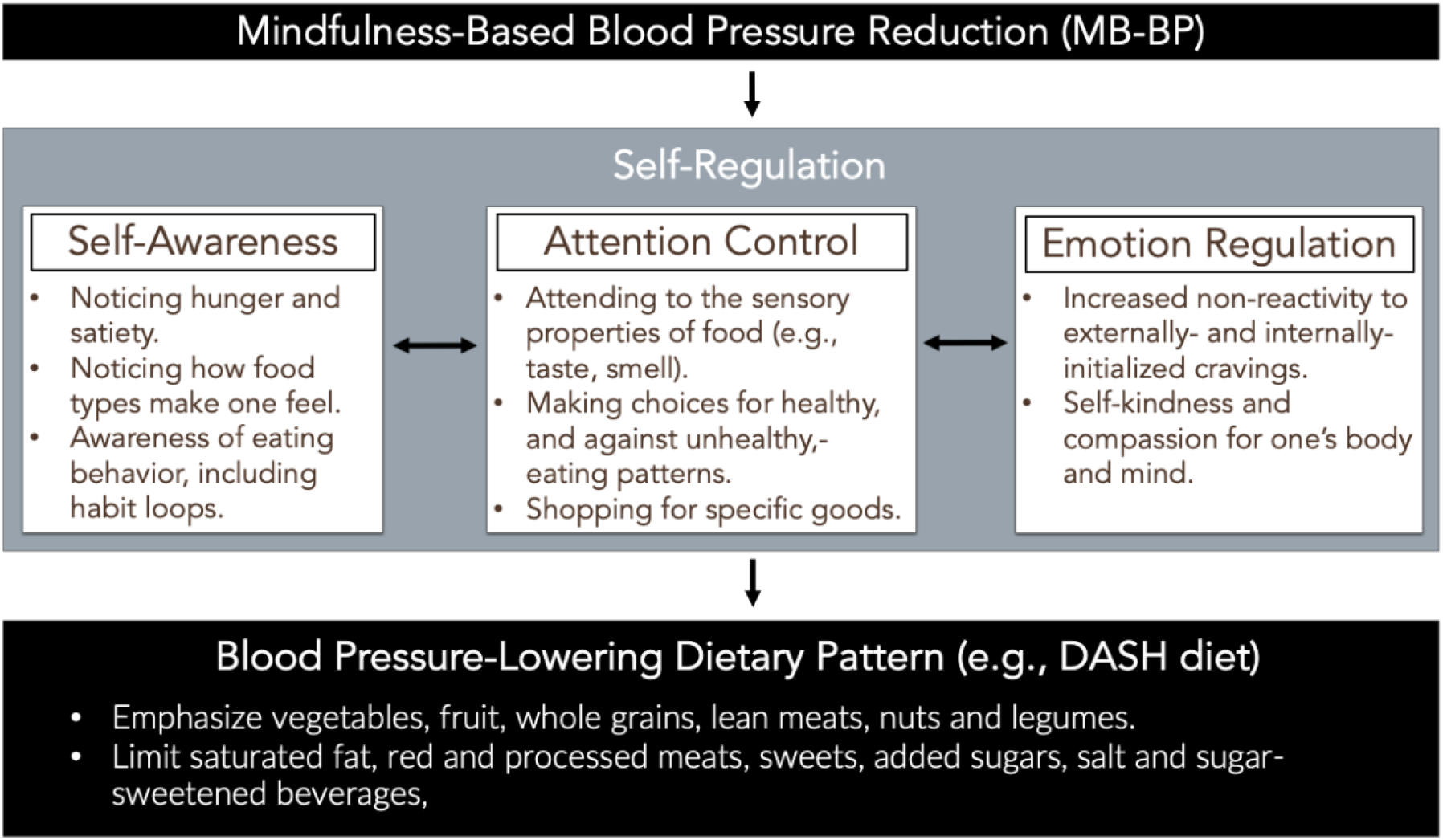
Theoretical framework of mechanisms through which Mindftilncss-Bascd Blood Pressure Reduction (MB-BP) may influence dietary patterns that lower blood pressure, such as the Dietary Approaches to Stop Hypertension (DASH) dietary pattern.

Participants assigned to the enhanced usual care control group received the same home BP device, training in home BP monitoring, and option for referral to primary care as participants in the intervention group. They also received an educational brochure about high BP and how to treat it, publicly available from the American Heart Association (product code 50-1731). Control group participants did not receive any mindfulness training as part of the study.

### Outcomes

The pre-specified primary outcome was interoceptive awareness, measured using the validated Multidimensional Assessment of Interoceptive Awareness (MAIA) at six months follow-up from baseline assessment and randomization.^17,18^ While the MAIA has eight dimensions, each with a separate scale, we utilized an eight-dimension average score (range 0 to 5) across the eight scales (i.e., mean score of the eight scales) to avoid issues of multiple statistical testing, similar to prior research.^9,19^ Secondary analyses evaluated the effect of the intervention on a six-dimension average MAIA score shown in factor analyses to represent one construct,^18,19^ and on each of the eight MAIA dimensions. Details are in **Supplement 2**.

Extent of adherence to the *Dietary Approaches to Stop Hypertension (DASH)-diet* was assessed using the Harvard 163-item 2007 Grid Food Frequency Questionnaire,^20^ and coding of DASH diet adherence using methods developed by Folsom et al.^21^ Specifically, the components of the DASH diet index were weighted and summed to calculate a single dietary concordance score (range 0-11). Details are in **Supplement 2**.

Key secondary outcomes that could be modifiable determinants of BP included measures of health behaviors, antihypertensive medication use, perceived stress, and mindfulness. *Physical activity* during the previous week was measured using the International Physical Activity Questionnaire (IPAQ) as total MET-minutes per week of physical activity, and time spent in sedentary sitting activities, with validation described elsewhere.^22^ Body mass index (kg/m^2^) was calculated by weight and height measures obtained from participants wearing light clothing without shoes, using a calibrated stadiometer (SECA, Hamburg, Germany) and weighing scale (SECA, Model 22089, Hamburg, Germany) operated by trained technicians. *Alcohol consumption* was assessed via a modified Centers for Disease Control and Prevention Behavioral Factor Surveillance System Questionnaire which has demonstrated concurrent validity with other nationally representative survey measures (e.g. NHIS, NHANES) in multiple studies evaluating alcohol consumption.^23^ *Stress* was assessed utilizing the 10-item Perceived Stress Scale (PSS-10) with established validity and reliability.^24^ *Emotional eating* was assessed using the Three Factor Eating Questionnaire Revised 21-item (TFEQ-R21).^25^ Mindfulness levels were assessed using the validated Five-Facet Mindfulness Questionnaire (FFMQ) a 39-item scale with established validity and reliability.^26^ Age, gender, race/ethnicity, and educational attainment were assessed using self-report.

We systematically measured adverse events, including monitoring serious adverse events and physical injuries through at-minimum-monthly online safety checks. All adverse events were documented and reported in accordance with the data safety monitoring plan, including reporting all serious adverse events to the study PI and the data safety monitoring board chair, detailed elsewhere.^15^

### Statistical Analysis

For statistical power considerations, another RCT of a mindfulness-based programs similar to MB-BP, specifically Mindfulness-Based College (n= 77), demonstrated a between-group effect size of 0.63 on the MAIA scale (SD=0.83).^27^ Conservatively, power analyses using the T statistic and non-centrality parameter, with alpha (two-tailed) set at 0.05 and beta at 0.2 with a MAIA difference of 0.40 or 0.50 and SD of 0.83 show sample size requirements of 68 or 45 per group, respectively. With approximately 100 participants in each group, the power was adequate assuming MAIA effect sizes of ≥0.40, with an anticipated dropout rate of 20%. For mediation analyses, Fritz and MacKinnon estimated that to detect a mediated effect when the effect of the exposure on the mediator and the mediator on the outcome was relatively small (standardized effect sizes of 0.14-0.39 each), using a delta-method based test as described below, a sample size of 422 was needed to obtain 80% power.^28^ As such, the mediation analyses reported in this paper are exploratory and underpowered.

We used generalized estimating equations (GEE) to evaluate the effect of MB-BP on the outcomes compared to the control at 3 and 6 months using an identity link and autoregressive covariance structure. Unadjusted analyses were prespecified, as there are risks for covariate adjustment making the models less precise and inducing bias.^29^ However, we also recognize recommendations to perform analyses adjusted for the strata used to randomize participants in (i.e., gender and BP status) in order to increase statistical power.^29^ As such, we present analyses adjusting for the randomization strata variables, while providing sensitivity analyses on the unadjusted models, so that readers have access to findings from both approaches. As some participants had healthy DASH-consistent diets at baseline and did not need to improve their diet, analyses on DASH diet in addition to looking at the full sample, also restricted the sample to those with poor DASH diet scores using the a priori defined cut-point of <5.5, as used elsewhere.^9,21^

Mediation analyses used formal mediation methods based on the counterfactual framework, which allows for decomposition of a total effect into direct and indirect effects, even in models with interactions and nonlinearities.^30^ Examining indirect effects provides evidence of whether mindfulness may exert its effects uniquely through plausible mediators examined in this study.

Analyses were performed as intention-to-treat (ITT). We analyzed all participants with data regardless of whether they completed the MB-BP or control programs. For example, of the 25 participants that had at least 2 absences from the MB-BP program, 13 (52%) took part in the 6-month follow-up assessments and were included in analyses. Of the 15 participants that withdrew/discontinued MB-BP, five took part in the 6-month follow-up assessments and were included in analyses. This complete case analysis is the main outcome analysis. We performed sensitivity analyses using multiple imputation. Predictive mean matching based on 14 baseline variables was used to impute missing SBP values at 6 months (e.g., mean SBP, mean DBP, PSS-10 score, FFMQ score, IPAQ, DASH diet score, BMI, age, race/ethnicity, and gender).^31^ Fifty imputations were generated and GEE analysis was performed in each. All analyses were performed using SAS version 9.4 (SAS Inc., Cary, NC), except for multiple imputation that was performed in R version 4.2.2 using mice package 1.

## Results

The sample was 58.7% female, 81.1% non-Hispanic white, 51.7% with a college degree, and an average age of 59.5 years (range: 22-84). There were no baseline differences between groups in demographics, MAIA score, DASH diet score, nor other behavioral determinants of BP (**Table 1**).

**Table 1.**
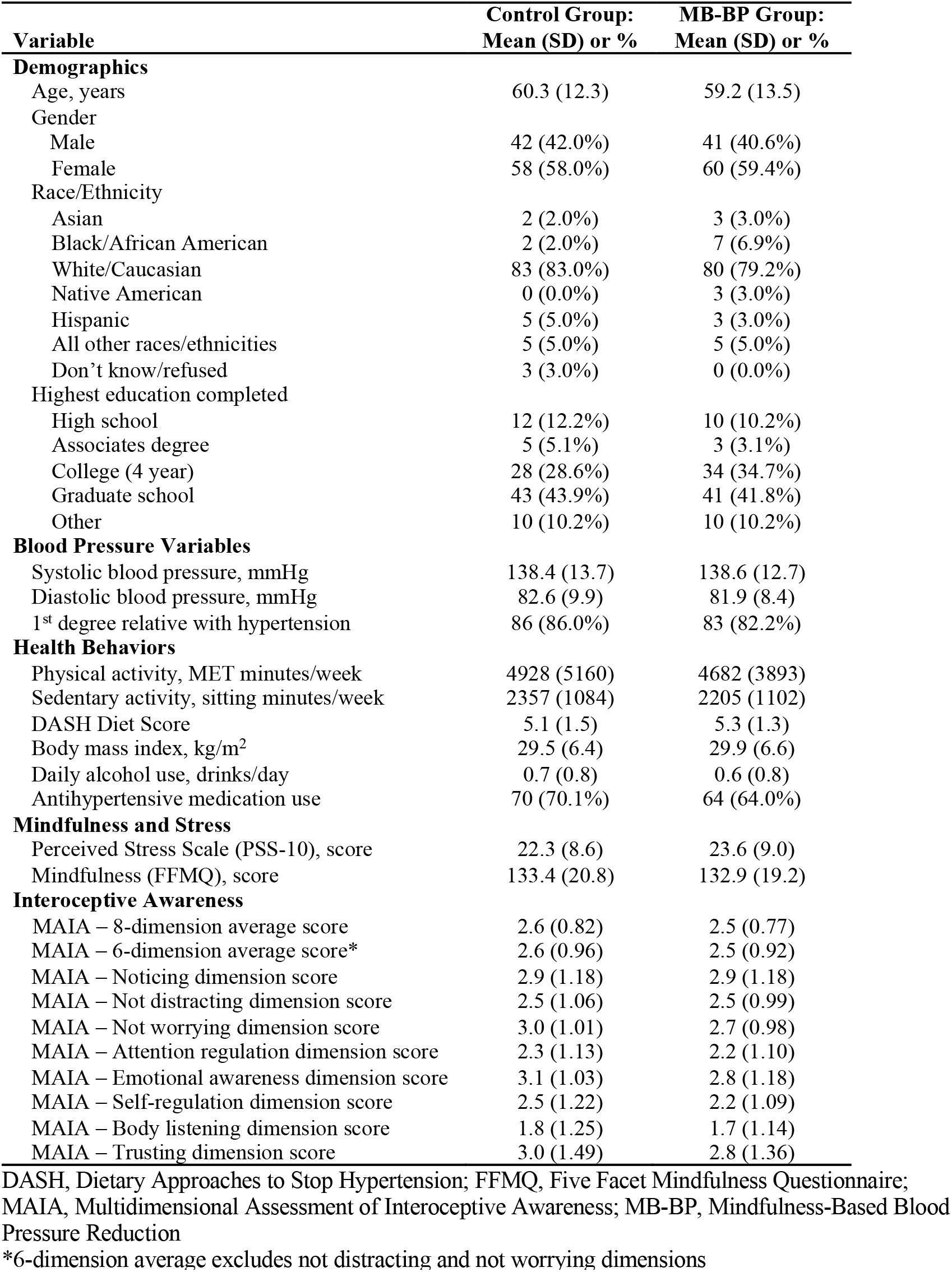
Baseline characteristics table for MB-BP group (n=101) and control group (n=100).

The CONSORT flow diagram is shown in **Figure 2**. There were 201 participants randomized to either MB-BP (n=101) or the enhanced usual-care group (n=100). Ninety-nine of the 101 participants assigned to MB-BP received the intervention, while all 100 assigned to the control group were provided the enhanced usual care components. Eighty-four of the 101 MB-BP participants (83.2%) attended at least seven of the ten MB-BP class sessions. The most common reason for discontinuing the intervention was health problems unrelated to the course (n=5); details are in **Figure 2**. Loss-to-follow-up rates across the two groups were similar with 12.8% (n=13) of MB-BP participants discontinuing the study vs. 11.0% (n=11) of control group participants. With the research study occurring during the years 2017 to 2020 and the COVID pandemic becoming a high-risk event in the USA shortly after the last round of enrollment (February/March 2020), all participants completed baseline assessments and began MB-BP prior to COVID. However, the pandemic forced a pivot to online delivery of MB-BP. The MB-BP intervention was taught by three certified instructors, teaching 41, 33, and 27 participants, respectively.

**Figure 2.**
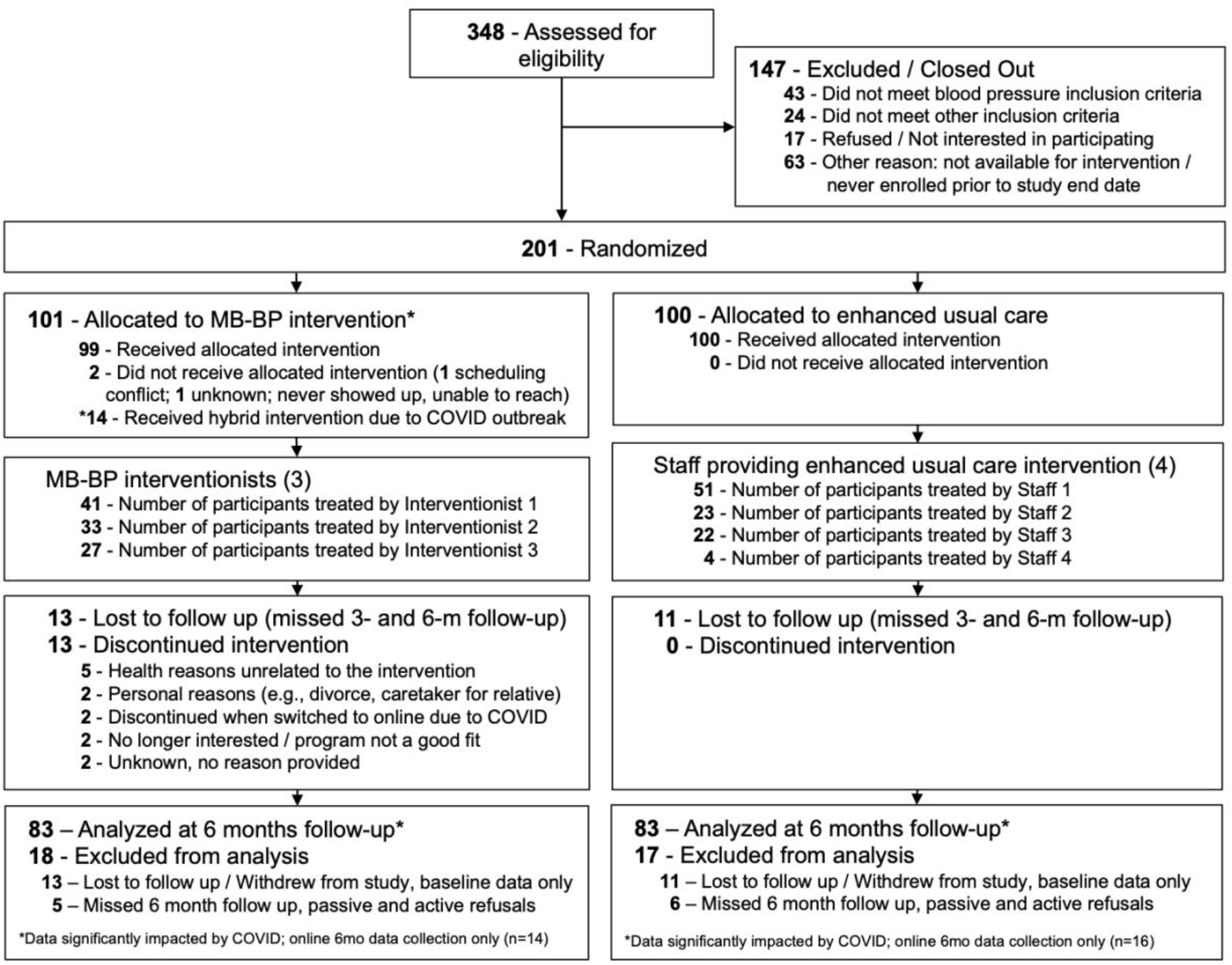
CONSORT flow diagram for participants randomized to MB-BP or enhanced usual care control.

Primary outcome analyses showed a 0.71 point improvement in the average interoceptive awareness MAIA score for the MB-BP group at 6 months follow-up (95% CI: 0.63, 0.88; p<.0001) compared to baseline. Regression analyses demonstrated a between-group difference of 0.54 (95% CI: 0.353, 0.735; p<.0001) points on the MAIA score at 6 months follow-up (**Figure 3**). Results for either unadjusted analysis (between-group difference of 0.54; 95% CI: 0.34, 0.74; p<0.0001), or multiple imputation (between-group difference of 0.49; 95% CI: 0.25, 0.72; p<0.001) showed similar between-group differences. Further analyses that focused only on the UH3 cohort (n=119) showed similar findings (between-group difference of 0.64; 95% CI: 0.38, 0.90; p<0.0001). Secondary analyses showed effects of MB-BP vs. control on six of the eight MAIA dimensions, specifically “noticing,” “attention regulation,” “emotional awareness,” “self-regulation,” “body listening” and “trusting” (**Table 2**).

**Figure 3.**
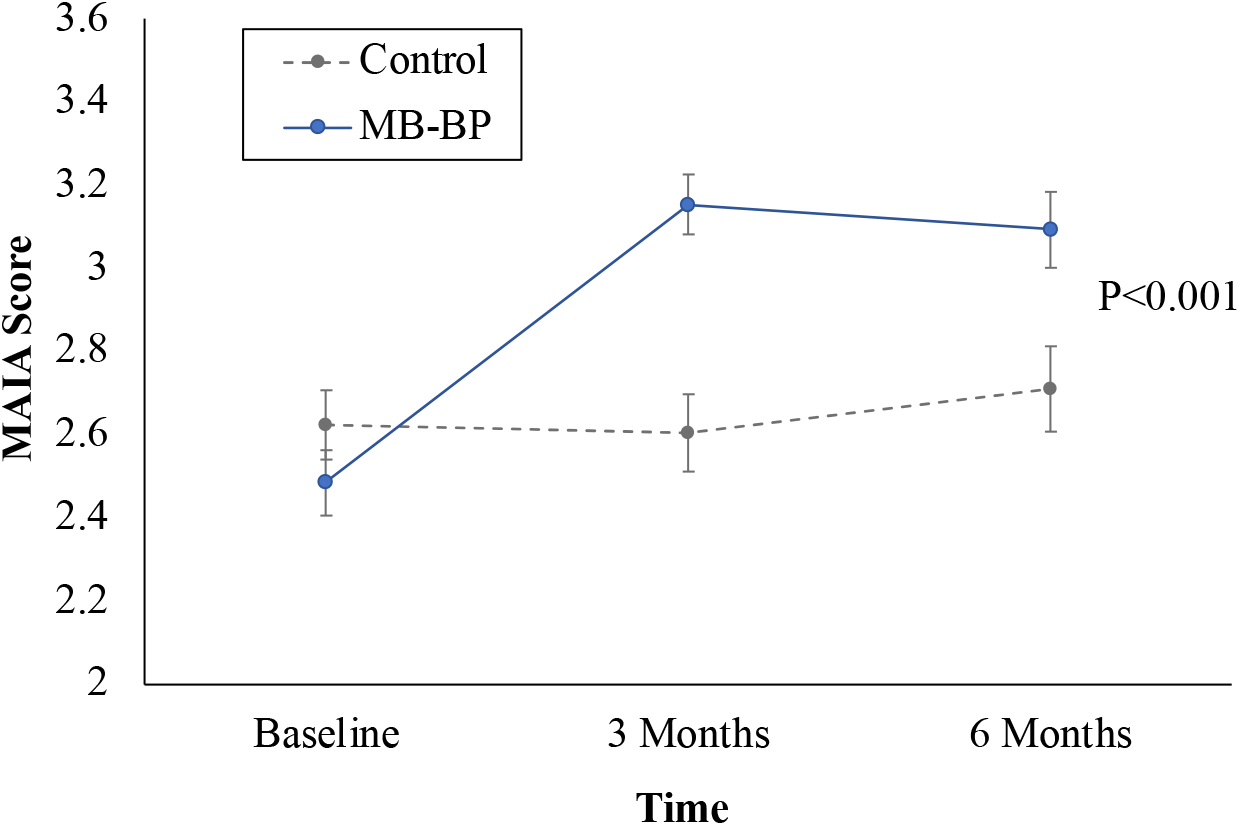
Impacts of MB-BP vs. control on interoceptive awareness, assessed using the Multidimensional Assessment of Interoceptive Awareness (MAIA). Error bars represent standard error of the mean.

**Table 2.** Between-group differences in change of mean Multidimensional Assessment of Interoceptive Awareness (MAIA) scores from baseline to follow-up, for MB-BP vs. contro groups.

| Outcome | Follow-Up Time |  |  |  |
| --- | --- | --- | --- | --- |
|  | 3 Months |  | 6 Months |  |
|  | Estimate | P | Estimate | P |
| MAIA 8-dimension average | 0.68 | <.0001 | 0.55 | <.0001 |
| MAIA 6-dimension average* | 0.78 | <.0001 | 0.65 | <.0001 |
| Dimension: Noticing | 0.35 | 0.02 | 0.36 | 0.03 |
| Dimension: Not Distracting | 0.36 | 0.02 | 0.13 | 0.44 |
| Dimension: Not Worrying | 0.19 | 0.15 | 0.10 | 0.49 |
| Dimension: Attention Regulation | 0.75 | <.0001 | 0.60 | <.0001 |
| Dimension: Emotional Awareness | 0.86 | <.0001 | 0.75 | <.0001 |
| Dimension: Self-Regulation | 1.03 | <.0001 | 0.79 | <.0001 |
| Dimension: Body Listening | 1.08 | <.0001 | 0.91 | <.0001 |
| Dimension: Trusting | 0.55 | 0.0007 | 0.48 | 0.003 |

\*6-dimension average excludes not distracting and not worrying dimensions
MB-BP, Mindfulness-Based Blood Pressure Reduction

The intervention was associated with a 0.34 point improvement in the DASH diet score in MB-BP participants from baseline (95% CI: 0.09, 0.59; p=0.01), while the control group showed a -0.04 point change in DASH diet score from baseline to 6 months follow-up (95% CI: - 0.31, 0.24; p=0.78; **Figure 4A**). Regression analyses showed a between-group difference of 0.32 (95% CI: -0.04, 0.68; p=0.08) at 6 months follow-up. Restricting the sample to the 97 participants with poor DASH diet scores (scores <5.5) at baseline demonstrated a 0.86 point (95% CI: 0.53, 1.19; p<.0001) improvement in the DASH diet score in the MB-BP group compared to baseline, and a 0.31 point (95% CI: -0.10, 0.71; p=0.13) change in the control group compared to baseline. Regression analyses demonstrated a between-group difference of 0.62 (95% CI: 0.13, 1.11; p=0.01) in MB-BP vs. control at 6-months follow-up (**Figure 4B**). Results for either unadjusted models (between-group difference of 0.63; 95% CI: 0.13, 1.12; p=0.01), or multiple imputation analysis showed similar findings (between-group difference of 0.59; 95% CI: 0.02, 1.15; p=0.04).

**Figure 4.**
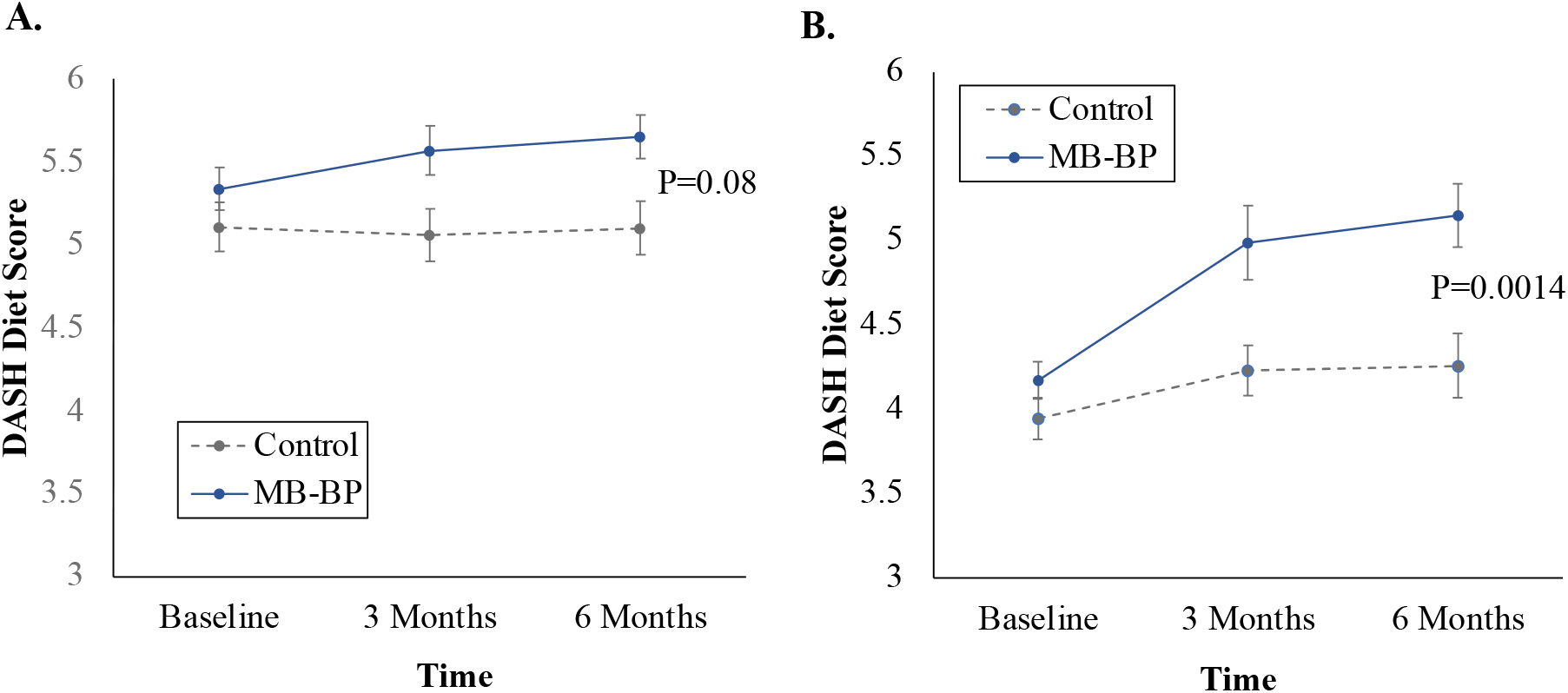
Impacts of MB-BP vs. control on change in Dietary Approaches to Stop Hypertension (DASH) diet score among (A) all participants, and (B) participants with poor DASH adherence at baseline (DASH score < 5.5; n=97). Error bars represent standard error of the mean.

In exploratory underpowered mediation analyses to evaluate whether MAIA may mediate effects of MB-BP on the DASH diet score, findings demonstrated partial mediation (31% mediated; p=0.28) for the MAIA 8-dimension average score (**Supplement 3**). Other plausible mediators that demonstrated evidence of mediation included sedentary behavior (sitting minutes per week; 12.6% mediated (p=0.25)) and mindfulness (33.1% mediated; p=0.23), shown in **Supplement 3**. Overall, findings suggest that interoceptive awareness may in part mediate the effects of MB-BP on DASH diet, as may sedentary behavior and mindfulness. Replication is required with larger sample sizes and greater statistical power.

There were eight serious adverse events observed, with four in the control group and four in the MB-BP group, during the 6 months of follow-up. Adverse events from physical injuries were also equal across the study groups (n=8 per group). None of the serious adverse events or physical injuries were found to be related to study involvement. More details on adverse events are reported elsewhere.^15^

## Discussion

Overall, findings show that MB-BP significantly improves interoceptive awareness compared to an enhanced usual care control group through 6 months of follow-up. Further, there was evidence that MB-BP improved the DASH dietary pattern, with larger effects in participants with poor adherence to the DASH dietary pattern at baseline.

In comparing this study’s findings on interoceptive awareness to other studies, we found impacts of a mindfulness training intervention on the MAIA, which is similar to other reports.^9,32^ One notable finding was that six of the MAIA domains (i.e., “noticing,” “attention regulation,” “emotional awareness,” “self-regulation,” “body listening” and “trusting” were significantly engaged and two (“not distracting”, “not worrying”) were not. A recent confirmatory factor analysis study suggested that the MAIA may not represent eight domains, but instead one, where all six domains we engaged with may represent one overarching domain of interoception, while the two domains we did not engage with may represent other constructs.^18,19^ Our findings are consistent with MB-BP influencing the overall construct of interoception.

Impacts of a mindfulness program on the DASH diet has only been assessed in one prior RCT to our knowledge, specifically, an underpowered pilot RCT in African American participants (n=25) with hypertension and mild cognitive impairment.^9^ The current study is the first adequately powered RCT we are aware of evaluating effects of mindfulness training on the DASH diet, and the first that has performed mediation analyses to evaluate whether mindfulness training-induced changes in self-awareness, specifically interoceptive awareness, translate into changes in dietary patterns. This study suggested that MB-BP improves DASH adherence, particularly in participants who have poor DASH adherence at baseline.

With regard to mechanisms, a theoretical framework for how mindfulness training could influence the DASH diet is emerging, described in more detail elsewhere^9,33^ and shown in **Figure 1**. Mechanisms by which mindfulness training can influence dietary behaviors can be categorized into three domains, specifically *self-awareness, attention control*, and *emotion regulation*, following the theoretical framework developed by Tang, Hölzel and colleagues and adapted to CVD risk reduction by Loucks and colleagues.^10^ In particular, *self-awareness* (of which interoceptive awareness is a component) can influence dietary behaviors, such as via (1) noticing hunger and satiety,^13^ (2) noticing how different food types make us feel,^6^ and (3) being aware of eating behaviors,^34^ including habits and reward-based learning.^35,36^ *Attention control* can be applied to dietary behaviors such as via (1) attending to the sensory properties of food (e.g., taste, smell, sight),^37^ (2) making conscious choices for healthy eating patterns and against possibly unconscious unhealthy eating patterns,^38^ and (3) shopping for specific (e.g., health-promoting) foods.^38^ Finally, *emotion regulation* can be applied to dietary behaviors such as via (1) decreased reactivity to externally- and internally-initialized cravings,^38^ and (2) self-kindness and compassion for one’s body and mind.^34^ Recent mechanistic findings from the MB-BP study, using diffusion tensor magnetic resonance imaging, demonstrated regions of the brain typically damaged by hypertension had either significant greater connectivity via improved mean diffusivity in the right fornix or significantly lessened damage measured by lower radial diffusivity in the left cingulum in MB-BP compared to control.^39^ The improvements in these regions were significantly correlated with interoceptive awareness measured via the MAIA, further supporting the role of interception in the effect of MB-BP on hypertension-related well-being.^39^ Indeed, focus groups and in-depth interviews of MB-BP participants suggested an ordering of mechanisms, where self-awareness often came first, following by strategies to regulate attention or emotion regulation.^33^

This study had several limitations. The follow-up time was 6 months; the durability of intervention effects are not known, although our prior single-arm study of the MB-BP intervention demonstrated significant pre-post improvements in interoceptive awareness and DASH diet through one year follow-up.^9^ The high proportion of well-educated white participants reduced generalizability, but this program may be a viable way of introducing health education in underserved populations in a way more specific to BP than MBSR alone, described by our group elsewhere.^15,40,41^ Third, this study meta-analyzed data across two previously unanalyzed clinical trials that pre-registered three primary outcomes (SBP, DASH diet score, and MAIA). The SBP outcome is reported elsewhere,^15^ and this manuscript reports on the other two outcomes in detail. Recognizing a Bonferroni correction for three outcomes would lower the p-value threshold to 0.017 for statistical significance, the between-group comparison p values for the MAIA in all participants (p<0.001) and DASH diet in those with poor diet at baseline (p=0.0014) are below those thresholds. Fourth, using the current study design, it was not possible to determine which components of MB-BP were most important for improving diet and interoceptive awareness. Future studies with dismantling or optimization designs can answer these questions. Finally, the study was underpowered to test mediation, which is important direction for future research so we can identify opportunities to optimize MB-BP.

### Perspectives

This study offers novel evidence that an adapted mindfulness training for participants with elevated BP that targets diet and interoceptive awareness improves both. Given the high burden of hypertension on CVD, the MB-BP program may offer a novel approach to improve adherence to evidence-based determinants of BP such as the DASH dietary pattern, in part through helping participants improve self-awareness of internal body sensations. These findings provide mechanistic evidence on how MB-BP lowers blood pressure.^9,15^

## Data Availability

In order to minimize the possibility of unintentionally sharing information that can be used to re-identify private information, a subset of the data generated for this study will be available at the Open Science Framework, accessed at doi.org/10.17605/OSF.IO/86UCD.

## Novelty and Relevance

1. What Is New?
  - This study shows that adapted mindfulness training improves interoceptive awareness and adherence to a blood pressure-lowering diet (i.e., Dietary Approaches to Stop Hypertension (DASH)).
2. What Is Relevant?
  - The DASH diet is one of the strongest dietary predictors of blood pressure, but adherence to the DASH diet is typically low. Adapted mindfulness training may help.
3. Clinical/Pathophysiological Implications?
  - The Mindfulness-Based Blood Pressure Reduction (MB-BP) program may improve one of the primary drivers of hypertension: a healthy eating pattern.

